# Trauma Exposure and Mental Health in Ex-Servicewomen Compared with Civilian Women in the UK

**DOI:** 10.64898/2026.03.18.26348680

**Authors:** Alexandria Smith, Lynsay Ayer, Sharon A.M. Stevelink

## Abstract

**Background:** Exposure to trauma is associated with poor mental health, but little is known about how trauma profiles differ between ex-servicewomen and civilian women. Differences in trauma exposure may arise before, during, and after military service.

**Objective:** To characterise trauma profiles in ex-servicewomen and civilian women in the UK using separate latent class analyses, and to examine associations between trauma class membership and mental health outcomes within each group.

**Methods:** Data were drawn from the UK Biobank and stratified by serving status. Ex-servicewomen (n = 446) were compared with civilian women (n = 54,068). Within each group, sixteen lifetime traumatic experiences were assessed, and latent class analysis was applied to identify trauma profiles. Multinomial logistic regression examined associations between class membership and sociodemographic characteristics, and logistic regression assessed associations between trauma classes and mental health difficulties.

**Results:** Five trauma classes were identified for both ex-servicewomen and civilian women. Ex-servicewomen were less likely than civilians to belong to the low-trauma class (33.0% vs 62.8%) and reported higher exposure to childhood trauma and intimate partner violence. Among civilians, all trauma classes were associated with elevated odds of depression, anxiety, self-injurious thoughts and behaviours (SITB), as well as reduced meaning in life. Among ex-servicewomen, associations were less consistent; only severe cumulative trauma was linked to all adverse mental health outcomes, while other classes showed no differences in anxiety compared to ex-servicewomen with low trauma exposures.

**Conclusion:** Trauma profiles and their mental health correlates differ between ex-servicewomen and civilian women. These differences may reflect early life vulnerabilities, military experiences, and post-service exposures. Although ex-servicewomen reported higher levels of trauma, the associations between trauma classes and mental health were less pronounced than among civilians.

**Highlights:**

- Ex-servicewomen showed substantially higher prevalence of trauma exposure compared to civilian women, with the greatest differences in childhood adverse events and intimate partner violence
- Separate latent class analyses identified five distinct trauma profiles in both groups, with ex-servicewomen considerably less likely to belong to the low-trauma class than civilian women (33.0% vs 62.8%).
- The association between trauma exposure and mental health outcomes was less consistent among ex-servicewomen than civilian women, suggesting that military service may involve resilience factors that moderate the trauma–mental health relationship.

## Introduction

Experiencing trauma during childhood and adolescence is a well-established risk factor for later adverse mental and physical health outcomes (1). Developmental theories and a broad body of observational research have consistently demonstrated that exposure to adversity during childhood can disrupt neurodevelopment, impair emotional regulation, and lead to long-term consequences for both physical and psychological health (2, 3). Early experiences and environments are believed to fundamentally alter brain development and body systems in ways that predispose individuals to chronic disease, mental health problems, and maladaptive behaviours across the life course (2, 3). Military service, which typically begins in early adulthood, a formative stage of developmental consolidation, may further increase the risk of trauma exposure and subsequent mental health difficulties (4). Exposure to trauma may sensitize individuals to subsequent mental health difficulties such as depression, anxiety, PTSD and substance use (5). Additionally, research shows that the greater number and types of potentially traumatic events a person experiences in childhood, the higher the risk for later mental health difficulties and other adverse health outcomes (1, 6).

Latent class analysis (LCA) and other “person centered” analytic approaches enable researchers to examine empirically-derived profiles of trauma and victimization across domains (e.g., child abuse and neglect, witnessing community violence, interpersonal violence) (7). These approaches have revealed that distinct profiles (classes) or combinations of trauma types differ in their demographic characteristics and associations with mental health difficulties (8).

Evidence indicates that trauma exposure patterns differ by sex across the lifespan. Among adolescents, boys are typically characterized by exposure to traumatic separation and physical violence across multiple contexts, including the home, school, and community, whereas girls are disproportionately represented in classes defined by emotional abuse and high levels of polytraumatization (8). Youth exposed to multiple forms of trauma are at greater risk for adverse outcomes, including suicidal behaviour, internalizing and externalizing symptoms, post-traumatic stress, and increased alcohol and substance use. Similar sex-specific patterns have been observed in adulthood, with women more frequently reporting direct interpersonal trauma such as physical or sexual assault (9). In a UK based study, using the UK Biobank data, distinct trauma profiles by sex were also identified (10). women exhibited a broader range of trauma profiles - including childhood adversity, intimate partner violence, sexual violence, and polytrauma, whereas men were more often characterized by physical and emotional trauma or sexual violence. Across studies, trauma exposure, irrespective of sex, was consistently associated with increased risk of mental health difficulties (8–10).

In the UK, little is known about how exposure to trauma differs between female civilians and those with military service. Women in the Armed Forces may be particularly vulnerable, not only because of elevated rates of childhood adversity among those who enlist (11), but also due to unique occupational and interpersonal stressors during service (12–14). These include combat exposure, witnessing violence or death, and heightened risk of interpersonal trauma such as intimate partner violence, harassment, or sexual assault (15, 16).

On the other hand, military service may confer protective effects. There is evidence that military service may attenuate the negative effects of childhood adversity on homelessness and self-reported general health (17). Among U.S. ex-servicewomen, most agreed that military service provided substantial professional benefits and career opportunities. The economic stability offered by military life may foster resilience and enable upward mobility, particularly for those with a history of early adversity (18). However, few studies have examined profiles of trauma in ex-servicewomen compared to civilian women, the differences in demographic characteristics, and how trauma profiles may be associated with mental health difficulties. Clarifying trauma profiles could improve our understanding of how early adversity, military experience, and post-service mental health intersect.

For this study, we expanded upon the work of Yapp and colleagues (2021) and used the UK Biobank to identify ex-servicewomen and compare their lifetime trauma exposure to civilian women (10). We identified trauma profiles among ex-servicewomen and civilians separately using latent class analysis (LCA) and examined whether these trauma profiles were differentially associated with demographic characteristics (e.g., age, ethnicity, education, and areas level of deprivation) and with mental health difficulties, including depression, anxiety, posttraumatic stress disorder (PTSD), and suicidal thoughts and behaviours, between the two groups.

## 2. Methods

### Participants and procedure

Data for this study were obtained from the UK Biobank, a large-scale prospective cohort study comprising individuals residing in England, Scotland, and Wales. Between 2006 and 2010, approximately 500,000 participants aged 40 to 69 years were recruited across 23 assessment centres from 2006 to 2010 (19). At baseline, participants provided detailed sociodemographic and medical histories, underwent a series of physical assessments, and contributed biological samples. The UK Biobank is linked to participants’ electronic health records, including data on hospital inpatient admissions, primary care encounters, and cancer and mortality registries. Subsequent follow-up assessments have been conducted, including the 2015 Occupational History Module and the 2016 Mental Health Questionnaire (MHQ), both of which were incorporated into the present analysis. A total of 70 participants were excluded due to requests for study withdrawal.

### Measures

#### Identification of Military Service

Military service history was ascertained using data from the Occupational History Module, completed by 120,148 participants. Within this module, respondents reported details of their employment history, including both current and prior positions, along with corresponding start and end dates. Occupations were classified according to the 2010 Standard Occupational Classification (SOC) system (20). Military service was identified using SOC codes for Commissioned Officers (1171) and Non-Commissioned Officers and Other Ranks (3311) (21). Based on these classifications, 546 women with prior military service and 66,235 women without military service were identified.

#### Trauma

Traumatic experiences and mental health history were assessed using the UK Biobank Mental Health Questionnaire (MHQ). Of the eligible sample, 412 of 546 ex-servicewomen (81.7%) and 50,203 of 66,235 civilian women (81.6%) completed the MHQ, with no significant difference in response rates.

Sixteen potentially traumatic exposures were assessed using the MHQ trauma subsection, categorized into childhood, adulthood, and lifetime experiences. Childhood exposures were adapted from the Childhood Trauma Screener (CTS-5) and comprised five domains: emotional neglect, physical neglect, emotional abuse, physical abuse, and sexual abuse (22). Adulthood traumas, defined as events occurring since age 16, included insecure attachment, physical intimate partner violence, psychological abuse by an intimate partner, sexual assault by an intimate partner, and financial insecurity. Lifetime exposures encompassed serious accidents, witnessing sudden or violent death, experiencing serious illness, combat exposure, sexual violence, and victimization by violent crime. All trauma exposure items were dichotomized for analysis. These definitions are aligned with the trauma definitions used previously in defining trauma in the UK Biobank (10). The complete item descriptions and response distributions by prior military service status are presented in Table 1.

**Table 1:** Demographics and Mental Health among women by serving status.

| Variable | Civilian women<br>N=50,203 | Civilian women<br>% | Ex-servicewomen<br>N=412 | Ex-servicewomen<br>% | P-value |
| --- | --- | --- | --- | --- | --- |
| Age & Ethnicity |  |  |  |  |  |
| White | 49,036 | 97.9 | 408 | 99.5 | 0.024 |
| Age | 55.4 (7.5) |  | 55.5 (7.5) |  | 0.81 |
| Education |  |  |  |  | <0.001 |
| College/University | 24,879 | 49.7 | 124 | 30.2 |  |
| A levels/AS levels | 7,509 | 15.0 | 78 | 19.0 |  |
| O levels/GCSEs | 10,259 | 20.5 | 134 | 32.6 |  |
| CSEs/NVQ/HND/Prof | 4,984 | 10.0 | 48 | 11.7 |  |
| Other education | 2,412 | 4.8 | 27 | 6.6 |  |
| Townsend Quintiles (Deprivation) |  |  |  |  | 0.488 |
| Q1 (least deprived) | 10,071 | 20.1 | 71 | 17.3 |  |
| Q2 | 10,051 | 20.0 | 77 | 18.7 |  |
| Q3 | 9,986 | 19.9 | 86 | 20.9 |  |
| Q4 | 10,021 | 20.0 | 92 | 22.4 |  |
| Q5 (most deprived) | 10,019 | 20.0 | 85 | 20.7 |  |
| Alcohol Use (AUDIT-C Score) |  |  |  |  | 0.037 |
| No alcohol use (0) | 4,757 | 9.5 | 49 | 11.9 |  |
| Low (1–2) | 13,415 | 26.8 | 130 | 31.6 |  |
| Moderate (3–5) | 21,783 | 43.5 | 163 | 39.7 |  |
| High (6–7) | 5,576 | 11.1 | 35 | 8.5 |  |
| Severe (8–12) | 4,527 | 9.0 | 34 | 8.3 |  |
| Mental Health |  |  |  |  |  |
| PTSD | 617 | 1.2 | 5 | 1.2 | 0.977 |
| Anxiety | 10,130 | 20.2 | 76 | 18.5 | 0.383 |
| Depression | 18,863 | 37.6 | 179 | 43.5 | 0.014 |
| Ever thought life not worth living |  |  |  |  | 0.348 |
| Never | 32,792 | 65.6 | 255 | 62.2 |  |
| Once | 7,819 | 15.7 | 71 | 17.3 |  |
| More than once | 9,366 | 18.7 | 84 | 20.5 |  |
| Contemplated self-harm |  |  |  |  | 0.001 |
| Never | 41,575 | 83.0 | 323 | 78.6 |  |
| Once | 4,483 | 9.0 | 58 | 14.1 |  |
| More than once | 4,009 | 8.0 | 30 | 7.3 |  |
| Ever self-harmed | 2,599 | 5.2 | 28 | 6.8 | 0.136 |
| Ever attempted suicide | 1,276 | 2.9 | 14 | 4.1 | 0.207 |
| Belief life has meaning |  |  |  |  | 0.013 |
| Not at all | 661 | 1.3 | 10 | 2.5 |  |
| A little | 2,792 | 5.7 | 28 | 6.9 |  |
| Moderate amount | 13,279 | 26.5 | 91 | 22.4 |  |
| Very much | 26,182 | 52.2 | 215 | 52.8 |  |
| An extreme amount | 6,493 | 12.9 | 63 | 15.5 |  |

PTSD = Post-Traumatic Stress Disorder; AUDIT-C = Alcohol Use Disorders Identification Test – Consumption; CSE = Certificate of Secondary Education; NVQ = National Vocational Qualification; HND = Higher National Diploma; GCSE = General Certificate of Secondary Education; AS/A levels = Advanced Subsidiary/Advanced Level qualifications.

#### Mental Health

A history of depression, anxiety, and post-traumatic stress disorder (PTSD) were identified from one of the three sources: self-reported diagnoses, symptom-based assessments, and hospital inpatient records.

Self-reported diagnoses were collected at two time points, at baseline (2006-2010) and follow-up (2016) using standardized questionnaires with slight variations in the question format. At baseline assessment, participants responded to the question: "Has a doctor ever told you that you have had any of the following serious medical conditions?" The list of conditions included “depression”, “anxiety/panic attacks”, and “post-traumatic stress disorder”. Participants were also invited to complete the MHQ in 2016, which asked: "Have you been diagnosed with one or more of the following mental health problems by a professional, even if you don’t have it currently?" and could indicate “depression” or “anxiety, nerves, or generalized anxiety disorder”. PTSD was not included as an option in this questionnaire.

Symptom-based diagnoses were also assessed through the MHQ using validated items from the Composite International Diagnostic Interview Short Form (CIDI-SF). Depression was classified using CIDI-SF depression module, lifetime version (23). Anxiety was classified with the CIDI-SF, anxiety module, lifetime version (23). Although the CIDI-SF was designed for DSM-IV disorders, the core symptom criteria for depression and generalized anxiety remained largely unchanged in DSM-5.

Using hospital inpatient records, mental health diagnoses were identified by *International Classification of Diseases 10th Revision (*ICD-10) diagnostic codes recorded in inpatient hospital data (24). The codes used were F32–F33 for depression, F40–F41 for anxiety disorders, and F43.1 for PTSD. Alcohol use was defined using Alcohol Use Disorders Identification Test (AUDIT-C), with three questions (1) How often did you have a drink containing alcohol in the past year? never (0 points); monthly or less (1 point); 2–4 times a month (2 points); 2–3 times a week (3 points); 4 or more times a week (4 points) (2) How many drinks containing alcohol did you have on a typical day when you were drinking in the past year? 1–2drinks(0points);3–4 drinks (1 point);5–6 drinks (2 points); 7–9 drinks (3 points); 10 or more drinks (4 points) and (3) How often did you have 6 or more drinks on one occasion in the past year? never (0 points); less than monthly (1 point);monthly (2 points); weekly (3 points); daily or almost daily (4 points). Scores were summed and categorised into five levels: no alcohol use (score = 0), low (1–2), moderate (3–5), high (6–7), and severe (8–12) (25).

Suicidal ideation, suicidal behaviours, and perceived meaning in life were assessed using the following questions: *“Many people have thoughts that life is not worth living. Have you felt that way?”* (No, yes once, yes more than once); *“Have you contemplated harming yourself (for example by cutting, biting, hitting yourself or taking an overdose)?”* (No, yes once, yes more than once); *“Have you deliberately harmed yourself, whether or not you meant to end your life?”* (Yes, no); and *“Have you harmed yourself with the intention to end your life?”* (Yes, no). Perceived meaning in life was assessed with the question: *“To what extent do you feel your life to be meaningful?”* Responses were categorised into three levels: high (an extreme amount, very much), moderate (a moderate amount), and low (a little, not at all).

Full variable definitions, coding criteria, and classification thresholds are provided in the supplemental tables.

#### Sociodemographic Variables

Sociodemographic variables included age at baseline assessment, sex, ethnicity (white/white British vs. other), and educational attainment (degree, A-levels, O-levels, or other). Area-level deprivation was assessed using the 2010 English Indices of Deprivation, a composite measure spanning income, employment, health, education, and environmental factors (26).

### Data analysis

To determine the optimal number of latent classes among ex-servicewomen and civilian women, we evaluated models with one to six classes, stratifying the sample by prior serving status. For each group (ex-servicewomen and civilian women), we assessed model fit using the Bayesian Information Criterion (BIC), sample size-adjusted BIC (aBIC), Akaike Information Criterion (AIC), and the log-likelihood ratio test statistic. Final model selection was based on balancing model fit, entropy, and conceptual coherence.

Multinomial logistic regression was subsequently used to examine whether sociodemographic characteristics (age, ethnicity, and area-level deprivation), perceived meaning in life (none, moderate, high), and the belief that life was worth living were associated with latent trauma class membership among ex-servicewomen and civilian women, respectively. In both models, the low trauma class served as the reference category.

Finally, associations between trauma class membership and mental health difficulties, including depression, anxiety, suicidal ideation, suicidal behaviours, and perceived meaning in life were assessed using multinomial logistic regression. We used complete case analysis for testing associations as there was a low level of missing among our demographic and trauma variables (<3%). All results are presented as odds ratios with corresponding 95% confidence intervals, using a significance level of p = 0.05 (two-tailed).

## 3. Results

### Sample Characteristics

Participants included 412 ex-servicewomen and 50,203 civilian women who had responded to both the occupational module and the MHQ. Demographic and mental health characteristics are presented in Table 1. Compared to female civilians, a higher percentage of ex-servicewomen identified as White (99.5% vs. 97.9%, p = 0.024) and a lower percentage reported university-level education (30.2% vs. 49.7%, p < 0.001). However, there were no significant differences in socioeconomic status, as measured by the Townsend Index of area-level deprivation.

### Mental Health Outcomes

A higher percentage of ex-servicewomen reported either current or past history of depression compared to civilians (43.5% vs. 37.6%, p = 0.014), while no significant differences were observed for current or past history of anxiety (18.5% vs. 20.2%, p = 0.383) or PTSD (1.2% vs. 1.2%, p = 0.977). A greater percentage of ex-servicewomen reported having contemplated self-harm once (14.1% vs. 9.0%, p = 0.001), although no group differences in percentages of those who contemplated self-harm more than once (7.3% vs. 8.0%). There were no significant differences in the percentage reporting self-harming behaviours (6.8% vs. 5.2%, p = 0.136), suicide attempts (4.1% vs. 2.9%, p = 0.207), or thoughts that life was not worth living (37.8% vs. 34.4%, p = 0.348).

Regarding perceived meaning in life, a higher percentage of ex-servicewomen selected “not at all” (2.5% vs. 1.3%) or “a little” (6.9% vs. 5.7%) compared to civilians, while a slightly higher percentage of ex-servicewomen also reported “an extreme amount” (15.5% vs. 12.9%) compared to civilians (p = 0.013).

### Trauma Exposure

Distributions of trauma exposure are presented in Table 2. Ex-servicewomen reported higher levels of trauma exposure across the lifespan compared to civilian women, with particularly elevated rates of childhood and adult interpersonal trauma. In terms of childhood trauma, ex-servicewomen reported higher percentages of emotional neglect (35.2% vs. 21.7%), physical abuse (30.1% vs. 16.9%), emotional abuse (24.0% vs. 17.1%), and sexual abuse (14.6% vs. 11.1%) compared to civilians (all *p* < 0.001).

**Table 2:** Distribution of Trauma.

| Distribution of trauma by serving status among women in the UK Biobank |  | Civilian Women<br>N= 50,203 |  | Ex-servicewomen<br>N=412 |  | P-value |
| --- | --- | --- | --- | --- | --- | --- |
|  |  | N | % | N | % |  |
| Emotional childhood neglect | When I was growing up I felt loved (less than “often”) | 10,868 | 21.7 | 145 | 35.2 | <0.001 |
| Physical childhood abuse | When I was growing up people in my family hit me so hard that it left me with bruises or marks (more than “never”) | 8,468 | 16.9 | 124 | 30.1 | <0.001 |
| Emotional childhood abuse | When I was growing up I felt like someone in my family hated me (more than “never”) | 8,560 | 17.1 | 99 | 24.0 | <0.001 |
| Physical childhood neglect | When I was growing up there was someone to take me to the doctor if I needed it (less than “very often”) | 7,442 | 14.8 | 88 | 21.4 | <0.001 |
| Sexual childhood abuse | When I was growing up there was someone to take me to the doctor if I needed it (less than “very often”) | 5,547 | 11.1 | 60 | 14.6 | <0.001 |
| Relationship Insecurity | Since I was sixteen, I have been in a confiding relationship (less than “often”) | 15,127 | 30.1 | 145 | 35.2 | 0.026 |
| Physical intimate partner violence | Since I was sixteen, a partner or ex-partner deliberately hit me or used violence in any other way (more than “never”) | 7,774 | 15.5 | 90 | 21.8 | <0.001 |
| Psychological intimate partner violence | Since I was sixteen, a partner or ex-partner repeatedly belittled me to the extent that I felt worthless (more than “never”) | 14,811 | 29.5 | 143 | 34.7 | 0.021 |
| Sexual intimate partner violence | Since I was sixteen, a partner or ex-partner sexually interfered with me, or forced me to have sex against my wishes (more than “never”) | 4,678 | 9.3 | 51 | 12.4 | 0.033 |
| Financial insecurity | Since I was sixteen, there was money to pay the rent or mortgage when I needed it (less than “very often”) | 7,014 | 14.0 | 69 | 16.8 | 0.107 |
| Serious accidents | In your life have you been in a serious accident that you believed to be life-threatening at the time (“yes”) | 3,398 | 6.8 | 34 | 8.3 | 0.233 |
| Witnessing a death | In your life have you witnessed a sudden violent death (e.g. murder, suicide, aftermath of an accident) (“yes”) | 4,640 | 9.2 | 55 | 13.4 | 0.004 |
| Serious illness | In your life have you been diagnosed with a life-threatening illness (“yes”) | 7,572 | 15.1 | 71 | 17.2 | 0.225 |
| Experiencing combat | In your life have you been involved in combat or exposed to a war-zone (either in the military or as a civilian) (“yes”) | 670 | 1.3 | 84 | 20.4 | <0.001 |
| Sexual violence | In your life have you been a victim of a sexual assault, whether by a stranger or someone you knew (“yes”) | 10,432 | 20.8 | 120 | 29.1 | <0.001 |
| Physically violent crime | In your life have you been attacked, mugged, robbed, or been the victim of a physically violent crime (“yes”) | 7,281 | 14.5 | 59 | 14.3 | 0.919 |

Ex-servicewomen also reported greater exposure to intimate partner violence, including physical (21.8% vs. 15.5%, *p* < 0.001), psychological (34.7% vs. 29.5%, *p* = 0.020), and sexual violence (12.4% vs. 9.3%, *p* = 0.033). Lifetime exposure to sexual violence was elevated among ex-servicewomen but was also substantial among civilians (29.1% vs. 20.8%, *p* < 0.001). Exposure to combat or war was also higher among ex-servicewomen compared to civilian women (20.4 vs. 1.3%%, p < 0.001).

By contrast, similar percentages were observed across groups for serious accidents, serious illness, and victimization by violent crime, indicating that differences were concentrated in specific trauma domains rather than uniformly distributed across all 16 trauma types.

### Latent class analysis

Separate latent class analyses were conducted for ex-servicewomen and civilian women. Model fit statistics for these analyses are presented in Table 3. For both groups, model fit improved with the addition of more classes, as evidenced by consistent decreases in the Akaike Information Criterion (AIC), Bayesian Information Criterion (BIC), and sample size-adjusted BIC, along with significant log-likelihood ratio tests at each step.

**Table 3:** Latent class fit statistics.

| Fit Statistics Civilian Women |  |  |  |  |  |  |  |  |
| --- | --- | --- | --- | --- | --- | --- | --- | --- |
| Classes | LogLikelihood | AIC | BIC | aBIC | Entropy | G<sup>2</sup> | p-value | Parameters |
| 1 | -328,548 | 657,128 | 657,269 | 657,218 | NaN |  |  | 16 |
| 2 | -309,131 | 618,328 | 618,619 | 618,514 | 0.724407 | 38,834 | <0.001 | 33 |
| 3 | -305,598 | 611,295 | 611,737 | 611,578 | 0.7822 | 7,066 | <0.001 | 50 |
| 4 | -302,780 | 605,694 | 606,285 | 606,073 | 0.796569 | 5,635 | <0.001 | 67 |
| 5 | -300,554 | 601,275 | 602,016 | 601,749 | 0.772619 | 4,453 | <0.001 | 84 |
| 6 | -298,988 | 598,178 | 599,069 | 598,748 | 0.746549 | 3,131 | <0.001 | 101 |
| Fit Statistics Ex-servicewomen |  |  |  |  |  |  |  |  |
| Classes | LogLikelihood | AIC | BIC | aBIC | Entropy | G<sup>2</sup> | p-value | Parameters |
| 1 | -3315.83 | 6663.652 | 6727.988 | 6677.225 | NaN |  |  | 16 |
| 2 | -3125.54 | 6317.077 | 6449.771 | 6345.058 | 0.730946 | 380.57 | <0.001 | 33 |
| 3 | -3060.85 | 6221.696 | 6422.747 | 6264.091 | 0.78218 | 129.38 | <0.001 | 50 |
| 4 | -3020.64 | 6175.285 | 6444.694 | 6232.083 | 0.756871 | 80.41 | <0.001 | 67 |
| 5 | -2978.31 | 6124.626 | 6462.392 | 6195.836 | 0.797666 | 84.66 | <0.001 | 84 |
| 6 | -2951.9 | 6105.789 | 6511.912 | 6191.429 | 0.834192 | 52.84 | <0.001 | 101 |

AIC= Akaike Information Criterion; BIC = Bayesian Information Criterion; aBIC = sample size adjusted BIC; G<sup>2</sup> Likelihood Ratio Chi Square

Among female civilians, the highest entropy was observed in the four-class solution (0.80), with a slight decline in the five-class model (0.77). For ex-servicewomen, the highest entropy was found in the six-class solution (0.83), suggesting that trauma experiences among ex-servicewomen may be more heterogeneous and complex compared to civilian women. However, based on interpretability and meaningful separation between classes, the five-class model was selected as optimal for both groups. While class structures showed conceptual similarities, the specific patterns and prevalences differed between ex-servicewomen and civilian women

The five-class solution was chosen for slightly different reasons in each group. While the four-class model had the highest entropy among civilians, the five-class model better distinguished women with severe childhood trauma but lower levels of adult revictimization from those with persistent, lifelong poly-traumatization. Among ex-servicewomen, the five-class solution was preferred over the six-class model despite marginally lower entropy (0.80 vs. 0.83), as it provided greater conceptual clarity and more balanced class sizes.

Ex-servicewomen showed more distributed trauma exposure across classes. The largest group was *Low Trauma Exposure* (33.0%), followed by *Adult IPV & Combat* (27.9%), which uniquely featured high combat exposure alongside intimate partner violence. *Childhood Abuse and Neglect* (16.8%) was characterized by severe childhood neglect and physical abuse. Both *Severe Cumulative Trauma* (11.2%) and *Child Sexual Abuse and Adult Sexual Violence* (11.2%) showed extensive trauma histories, with the *Severe cumulative Trauma* including childhood emotional abuse and neglect in addition to adult IPV and combat exposure. *Child Sexual Abuse and Adult Sexual Violence* predominantly included sexual violence across the lifespan.

Civilian women demonstrated greater concentration in low-trauma categories compared to ex-servicewomen, with 62.8% in *Low Trauma Exposure*. The remaining classes were smaller: *Adult IPV* (11.5%), *Childhood Abuse and Neglect* (11.5%), *Child Sexual Abuse and Adult Sexual Violence* (8.9%), and *Severe Cumulative Trauma* (5.4%). This five-class structure is consistent with Yapp et al. 2023, who identified conceptually similar trauma class categories among females in the UK Biobank(10).

### Sociodemographic characteristics by trauma classification

Among ex-servicewomen, there were no major demographic differences across trauma classes compared to the low trauma group (Table 4), with age, ethnicity, education, and deprivation showing mostly non-significant or inconsistent associations. However, ex-servicewomen in all trauma classes were less likely to report that their lives had a great deal of meaning compared to those in the low trauma group.

**Table 4:** Multinomial logistic regression analysis of demographic characteristics by latent class groups in ex-servicewomen versus civilian women.

|  |  |  |  |  |  |  |  |  |
| --- | --- | --- | --- | --- | --- | --- | --- | --- |
| Ex-servicewomen demographics by latent classes |  |  |  |  | Civilian women demographics by latent classes |  |  |  |
| Reference: Low trauma | Adult IPV & combat | Child sexual abuse & adult sexual violence | Severe cumulative trauma | Childhood abuse & neglect | Adult IPV | Child sexual abuse & adult sexual violence | Severe cumulative trauma | Childhood abuse & neglect |
| Age | **0.94 (0.9-0.97)** | 0.98 (0.93-1.04) | 0.95 (0.9-1) | 1.02 (0.97-1.06) | **0.98 (0.98-0.98)** | **0.98 (0.97-0.98)** | **0.96 (0.95-0.96)** | **0.99 (0.99-0.99)** |
| White | 0.83 (0.38-1.79) | 0 (0-0) | 0.33 (0.08-1.35) | 0 (0-0) | **0.69 (0.57 – 0.85)** | **0.51 (0.42-0.62)** | **0.25 (0.21-0.31)** | **0.47 (0.39-0.56)** |
| Townsend level of deprivation quintiles (*Ref. least deprived*) |  |  |  |  |  |  |  |  |
| 2 | 0.48 (0.21-1.08) | 1.04 (0.25-4.34) | 1.71 (0.45-6.53) | 0.75 (0.22-2.51) | **1.19 (1.08-1.31)** | 1.06 (0.96-1.17) | 1.06 (0.92-1.23) | 1.08 (0.98-1.18) |
| 3 | 0.72 (0.32-1.62) | 3.23 (0.88-11.92) | 2.47 (0.63-9.63) | 2.49 (0.83-7.5) | **1.23 (1.12-1.35)** | 1.08 (0.97-1.19) | **1.33 (1.15-1.53)** | **1.16 (1.06-1.27)** |
| 4 | 0.52 (0.24-1.14) | 1.83 (0.5-6.73) | 1.28 (0.33-5.05) | 1.17 (0.39-3.52) | **1.50 (1.37-1.64)** | **1.30 (1.17-1.43)** | **1.88 (1.65-2.15)** | **1.29 (1.17-1.41)** |
| 5 | 0.85 (0.36-2.03) | 2.53 (0.67-9.61) | 2.08 (0.53-8.16) | 2.63 (0.89-7.76) | **1.94 (1.78-2.13)** | **1.67 (1.51-1.84)** | **3.08 (2.71-3.49)** | **1.54 (1.41-1.68)** |
| Education (*ref. Degree*) |  |  |  |  |  |  |  |  |
| A levels/AS levels | 0.98 (0.46-2.11) | 0.78 (0.28-2.17) | 1.95 (0.67-5.69) | 0.74 (0.28-1.98) | **1.10 (1.02-1.20)** | **0.85 (0.77-0.93)** | **0.89 (0.77-0.97)** | 1.02 (0.94-1.11) |
| O levels/GCSEs | 1.34 (0.69-2.61) | 1.05 (0.45-2.41) | 2.02 (0.78-5.19) | 0.73 (0.32-1.62) | 1.01 (0.94-1.09) | **0.77 (0.70-0.83)** | **0.87 (0.78-0.96)** | 1.00 (0.93-1.08) |
| CSEs/NVQ/HN D/Prof | 1.43 (0.59-3.49) | 0.14 (0.01-1.67) | 2.08 (0.59-7.32) | 1.31 (0.48-3.55) | 0.95 (0.86-1.05) | **0.82 (0.74-0.92)** | **1.15 (1.02-1.31)** | **1.26 (1.15-1.38)** |
| Other education | 0.13 (0.02-1.04) | 0.17 (0.02-1.67) | 1.31 (0.28-6.19) | 0.56 (0.17-1.82) | **0.75 (0.65-0.87)** | **0.52 (0.43-0.62)** | 0.86 (0.71-1.01) | **1.17 (1.04-1.33)** |
| Belief life has meaning (ref none/little) |  |  |  |  |  |  |  |  |
| Moderate amount | 0.35 (0.08-1.51) | **0.17 (0.04-0.86)** | 0.30 (0.07-1.32) | **0.16 (0.04-0.66)** | **0.64 (0.57-0.71)** | **0.67 (0.59-0.77)** | **0.36 (0.32-0.41)** | **0.47 (0.42-0.52)** |
| Very much | **0.3 (0.08-1.16)** | **0.12 (0.03-0.53)** | **0.07 (0.02-0.30)** | **0.07 (0.02-0.27)** | **0.41 (0.37-0.46)** | **0.49 (0.43-0.55)** | **0.15 (0.13-0.17)** | **0.23 (0.21-0.25)** |

PTSD = Post-Traumatic Stress Disorder; AUDIT-C = Alcohol Use Disorders Identification Test – Consumption; CSE = Certificate of Secondary Education; NVQ = National Vocational Qualification; HND = Higher National Diploma; GCSE = General Certificate of Secondary Education; AS/A levels = Advanced Subsidiary/Advanced Level qualifications.

**Table 5:** Multinomial logistic regression results for associations between mental health and suicidal thoughts and behaviours among ex-servicewomen versus civilian women.

| Ex-servicewomen |  |  |  |  | Civilian Women |  |  |  |
| --- | --- | --- | --- | --- | --- | --- | --- | --- |
| Reference: Low trauma | Adult IPV & combat | Child sexual abuse & adult sexual violence | Severe cumulative trauma | Childhood abuse & neglect | Adult IPV | Child sexual abuse & adult sexual violence | Severe cumulative trauma | Childhood abuse & neglect |
| Depression | **1.97 (1.15-3.4)** | 1.52 (0.73-3.18) | **2.73 (1.35-5.52)** | **2.45 (1.29-4.63)** | **2.33 (2.20-2.47)** | **2.04 (1.91-2.18)** | **5.28 (4.84-5.75)** | **2.37 (2.24-2.52)** |
| Anxiety | 1.53 (0.76-3.06) | 0.77 (0.28-2.14) | **2.65 (1.13-6.22)** | 1.96 (0.89-4.31) | **1.86 (1.74-1.99)** | **1.67 (1.55-1.80)** | **3.50 (3.22-3.81)** | **2.03 (1.90-2.17)** |
| Life not worth living | **2.82 (1.51-5.27)** | **3.64 (1.73-7.64)** | **11.45 (5.09-5.76)** | **4.24 (2.14-8.4)** | **2.85 (2.68-3.02)** | **2.4 (2.25-2.56)** | **7.09 (6.49-7.75)** | **3.18 (2.99-3.37)** |
| Thoughts of self-harm | **4.66 (1.87-11.63)** | **8.88 (3.11-25.37)** | **27.58(9.72-78.23)** | **7.25 (2.72-19.29)** | **3.24 (3.02-3.48)** | **2.97 (2.74-3.21)** | **8.71 (7.99-9.49)** | **3.20 (2.98-3.43)** |
| Ever self-harm | 4.88 (0.91-26.22) | 2.52 (0.28-22.60) | **16.78(3.08-91.32)** | **6.40(1.25-32.69)** | **3.95 (3.50-4.46)** | **3.44(3.01-3.94)** | **11.62(10.29-13.13)** | **3.84(3.40-4.35)** |
| Ever Suicide Attempt | 0.44 (0.03-6.00) | 1.25 (0.09-18.26) | **18.42 (2.6-130.4)** | **8.03 (1.25-51.71)** | **4.94 (4.14-5.90)** | **4.18 (3.43-5.10)** | **18.74 (15.84-22.16)** | **4.80 (4.02-5.74)** |
| Belief life has meaning* |  |  |  |  |  |  |  |  |
| Moderate meaning | 0.33 (0.08-1.47) | **0.18 (0.03-0.96)** | 0.29 (0.06-1.35) | **0.15 (0.03-0.7)** | **0.68 (0.60-0.76)** | **0.71 (0.62-0.81)** | **0.41 (0.36-0.47)** | **0.49 (0.44-0.54)** |
| A lot of meaning | 0.29 (0.07-1.17) | **0.15 (0.03-0.67)** | **0.07 (0.02-0.32)** | **0.07 (0.02-0.31)** | **0.45 (0.40-0.50)** | **0.52 (0.46-0.59)** | **0.17 (0.15-0.19)** | **0.24 (0.22-0.26)** |

Adjusted for age, ethnicity, education, area level indicator of deprivation (Townsend index), alcohol use (AUDIT-C)
\*Reference: Little or no meaning

Conversely, among civilian women, some sociodemographic factors were strongly associated with trauma class membership. Greater deprivation, non-White ethnicity, and lower educational attainment were associated with a higher odd of belonging to most trauma classes compared to low trauma. As with ex-servicewomen, low trauma was associated with a belief that life had meaning.

### Mental health outcomes and trauma classes

When examining mental health difficulties by trauma class, using the low trauma group as the reference, distinct profiles emerged between ex-servicewomen and civilian women. Among ex-servicewomen, depression was significantly associated with the *adult IPV & combat*, *severe cumulative trauma*, and *childhood abuse & neglect* classes. Anxiety was only significantly associated with the *severe cumulative trauma* class. All trauma classes were linked to greater odds of reporting that life was not worth living or having thoughts of self-harm. However, only the *Severe cumulative trauma* and *childhood abuse and neglect* classes were significantly associated with suicide attempts.

Among civilian women, a consistent pattern of elevated risk was observed across all trauma classes. Depression and anxiety were significantly associated with each class, with the *severe cumulative trauma* class showing the strongest associations. All trauma classes were also significantly associated with increased odds of reporting that life was not worth living, thoughts of self-harm, suicide attempts, and reduced belief that life had meaning, both at moderate and high levels compared to little/not at all.

## 4. Discussion

Ex-servicewomen experienced a higher cumulative burden of trauma than civilian women, with more diffuse and intersecting trauma profiles. In stratified latent class analyses, both groups independently yielded five conceptually similar classes, yet the distribution across classes differed substantially. Only one-third of ex-servicewomen belonged to the low-trauma class, compared with nearly two-thirds of civilian women. Among ex-servicewomen, trauma exposure was more evenly distributed across classes, with greater overlap between childhood adversity, intimate partner violence, and combat exposure, suggesting less discrete boundaries between trauma profiles in this population.

The pattern of associations between trauma class membership and mental health outcomes also appeared to differ between the two groups. Among civilian women, all trauma classes were consistently associated with elevated odds of depression, anxiety, and suicidal thoughts and behaviours relative to the low-trauma class, with the strongest associations observed in the severe cumulative trauma class. Among ex-servicewomen, a more heterogeneous pattern emerged. Depression and suicidal ideation were associated with multiple trauma classes, but anxiety was significantly associated only with the severe cumulative trauma class. These findings suggest different patterns of vulnerability and potentially protective factors among civilian women and ex-servicewomen.

### Trauma Exposure

Ex-servicewomen reported substantially higher prevalence of trauma compared to civilian women, with the starkest differences in childhood adversity; emotional neglect (35.2% vs. 21.7%), physical abuse (30.1% vs. 16.9%), emotional abuse (24.0% vs. 17.1%), and sexual abuse (14.6% vs. 11.1%). These findings are consistent with research documenting elevated childhood adversity among women in the AF compared to civilian women (27). Ex-servicewomen also reported greater exposure to intimate partner violence, including physical (21.8% vs. 15.5%), psychological (34.7% vs. 29.5%), and sexual IPV (12.4% vs. 9.3%). These IPV estimates are consistent with pooled estimates among women in military populations but exceed the lifetime IPV prevalence reported in the general female population of England and Wales in the British Crime Survey (28). By contrast, exposures not tied to childhood adversity, interpersonal contexts and combat exposure, such as serious accidents, serious illness, and violent crime, were similar across groups.

Beyond prevalence, the structure of trauma exposure also differed. Among ex-servicewomen, the latent class analysis revealed a unique Adult IPV & Combat class, comprising over a quarter of the sample, in which combat exposure co-occurred with intimate partner violence. Childhood abuse also frequently co-occurred with adult interpersonal trauma with adversity that spanned the life course. This pattern of intersecting childhood, interpersonal, and occupational trauma may reflect compounding pathways, wherein early adversity increases vulnerability to both military-specific exposures and post-service revictimization. In contrast, among civilian women, trauma classes were more discrete, with clearer separation between childhood-onset and adult-onset profiles. Most of the sample concentrated in the low-trauma class.

### Mental Health

The pattern of associations between trauma profiles and mental health outcomes differed markedly between ex-servicewomen and civilian women. Among civilian women, all trauma classes were associated with elevated odds of depression, anxiety, and suicidal thoughts and behaviours, with the strongest associations observed in the severe cumulative trauma class. In contrast, trauma class membership among ex-servicewomen showed a less consistent relationship with mental health outcomes, and wider confidence intervals, reflecting smaller sample sizes within classes. Among ex-servicewomen, severe cumulative trauma was associated with depression and suicidality relative to other trauma classes and was the only trauma class significantly associated with anxiety.

These findings may initially appear counterintuitive, as common mental disorders are more prevalent among UK serving and ex-serving personnel compared to civilians (29). Consistent with this literature, ex-servicewomen in our sample had a higher prevalence of depression and prior thoughts of self-harm, while prevalence of anxiety, PTSD, and suicidal behaviours was similar to civilian women. However, despite greater trauma exposure and higher baseline prevalence of some mental health conditions, the gradient between trauma class membership and mental health outcomes was less pronounced among ex-servicewomen than among civilian women. This distinction suggests that trauma exposure alone does not fully account for mental health outcomes in this population.

Several mechanisms may explain this divergence. Women who successfully complete military service may have higher distress tolerance, emotional regulation skills, or adaptive coping strategies, that buffer trauma’s impact on mental health outcomes (30). Military structure and community may also provide ongoing protective factors. The hierarchical organization, clear role expectations, social cohesion, and collective identity fostered in military environments can mitigate trauma’s psychological effects through unit cohesion mechanisms (31). Research demonstrates that military personnel with high unit cohesion show significantly lower odds of PTSD symptoms and psychological distress compared to those with low cohesion. Our findings demonstrate that the relationship between trauma and mental health is not uniform across populations, where a constellation of factors provide both risk and resilience among ex-servicewomen.

### Study Strengths and Limitations

This study benefits from a large population-based sample that allows for direct comparison between ex-servicewomen and civilian women drawn from the same cohort. However, several limitations should be noted. The smaller sample size of ex-servicewomen may have limited statistical power and reduced precision. However, the consistent pattern across outcomes and successful detection of other associations (e.g., depression) suggests these may represent genuine differences rather than solely reflecting power limitations. Lastly, the UK Biobank represents a more educated and predominantly white population, limiting generalizability to more diverse populations. Nevertheless, since both ex-servicewomen and civilian women were drawn from the same source population, between-group comparisons remain valid.

### Conclusion

Ex-servicewomen and civilian women show markedly different trauma profiles and mental health associations. Whether these differences stem from military service experiences, pre-existing characteristics that influence military enlistment, or both, remains unclear. Regardless of underlying causes, these distinct profiles have important implications for tailoring mental health services to ex-servicewomen populations.

**Figure 1:**
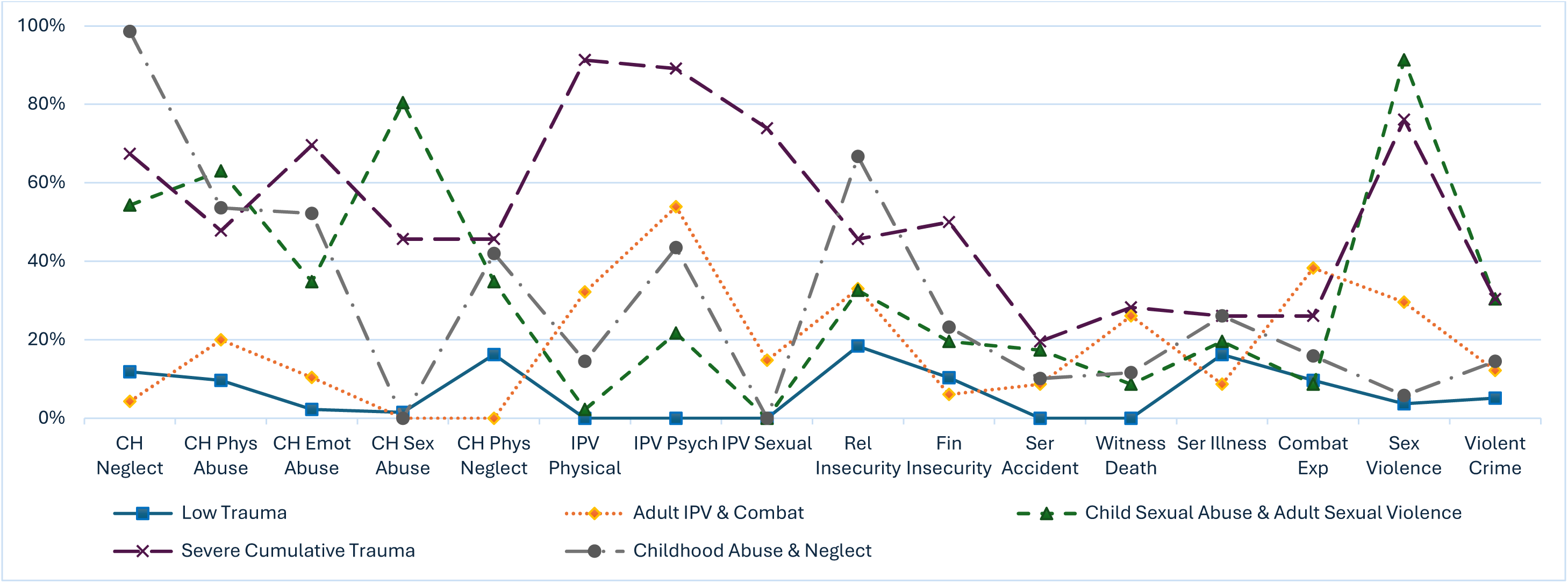
Five-class trauma distribution among ex-servicewomen. CH Neglect=emotional childhood neglect; CH Phys Abuse =physical childhood abuse; CH Emot Abuse=emotional childhood abuse; CH Sex Abuse = sexual childhood abuse; CH Phys Neglect = physical childhood neglect; IPV Physical =physical intimate partner violence; IPV Psych =psychological intimate partner violence; IPV Sexual =sexual intimate partner violence; Rel Insecurity=relationship insecurity; Fin Insecurity =financial insecurity; Ser Accident = serious accidents; Witness Death = witnessing a death; Ser Illness= serious illness; Combat Exp= experiencing combat; Sex Violence= sexual violence; Violent Crime = physically violent crime

**Figure 2:**
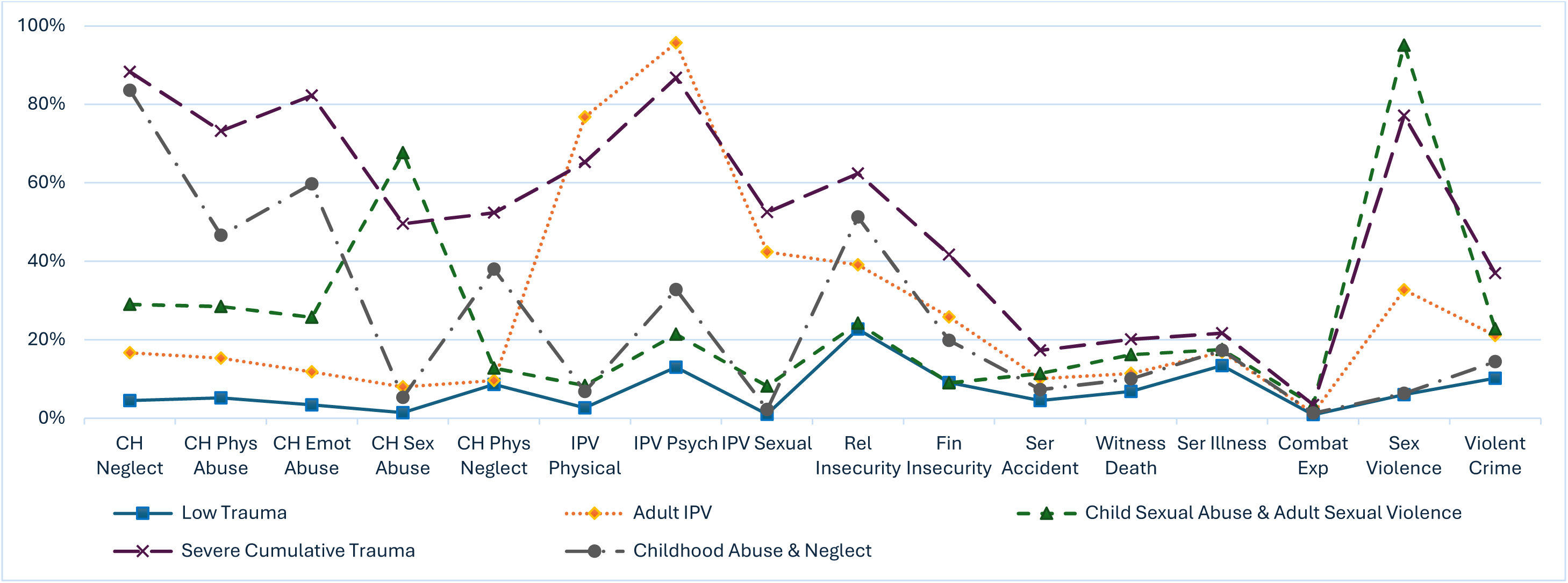
Five-class trauma distribution among civilian women. CH Neglect=emotional childhood neglect; CH Phys Abuse =physical childhood abuse; CH Emot Abuse=emotional childhood abuse; CH Sex Abuse = sexual childhood abuse; CH Phys Neglect = physical childhood neglect; IPV Physical =physical intimate partner violence; IPV Psych =psychological intimate partner violence; IPV Sexual =sexual intimate partner violence; Rel Insecurity=relationship insecurity; Fin Insecurity =financial insecurity; Ser Accident = serious accidents; Witness Death = witnessing a death; Ser Illness= serious illness; Combat Exp= experiencing combat; Sex Violence= sexual violence; Violent Crime = physically violent crime

## Acknowledgements

*This paper is dedicated to the memory of Professor Nicola T Fear, who sadly passed away in February 2026, as we finalised this manuscript. As a researcher, educator, and mentor, Nicola gave generously of her expertise, and we were privileged to work alongside her. She leaves behind a legacy of research that will improve the lives of the Armed Forces community for generations to come. We all loved you and miss you dearly.*

## Ethical Approval

Ethical approval for using UK Biobank was provided by the Northwest Multi-centre Research Ethics Committee (MREC) as a Research Tissue Bank (RTB).

## Declaration of interest

This research was partly supported by a grant from the Forces in Mind Trust (FiMT) FiMT/2202. No other potential conflicts of interest or financial disclosures were reported by the authors of this paper. SAMS is supported by the National Institute for Health and Care Research (NIHR) Maudsley Biomedical Research Centre at South London and Maudsley NHS Foundation Trust and funded by the National Institute for Health and Care Research, NIHR Advanced Fellowship, NIHR300592. The views expressed in this publication are those of the author(s) and not necessarily those of the NHS, the NIHR or the Department of Health and Social Care.

This research has been conducted using the UK Biobank Resource under Application Number 213421

## Data Availability Statement

The data used in this study are available from the UK Biobank (https://www.ukbiobank.ac.uk). Researchers can apply for access to UK Biobank data via the Access Management System at https://www.ukbiobank.ac.uk/enable-your-research/apply-for-access. Data cannot be made freely available due to participant confidentiality restrictions and data sharing agreements. This study used UK Biobank Application Number 213421.

## Author Contributions Statement

A.S. contributed to the conception and design of the study, data analysis and interpretation, drafting of the manuscript, and critically revised the manuscript. L.A. contributed to study design, data interpretation, and critically revised the manuscript S.S. contributed to data interpretation and critically revised the manuscript. All authors approved the final version to be published.

## References

1. Nelson CA, Bhutta ZA, Burke Harris N, Danese A, Samara M. Adversity in childhood is linked to mental and physical health throughout life. BMJ. 2020;371:m3048.

2. Felitti VJ, Anda RF, Nordenberg D, Williamson DF, Spitz AM, Edwards V, et al. Relationship of childhood abuse and household dysfunction to many of the leading causes of death in adults: the adverse childhood experiences (ACE) study. Am J Prev Med. 1998;14(4):245–58.

3. Shonkoff JP, Garner AS; Committee on Psychosocial Aspects of Child and Family Health; Committee on Early Childhood, Adoption, and Dependent Care; Section on Developmental and Behavioral Pediatrics. The lifelong effects of early childhood adversity and toxic stress. Pediatrics. 2012;129(1):e232–46.

4. Arnett JJ. Emerging adulthood: a theory of development from the late teens through the twenties. Am Psychol. 2000;55(5):469–80.

5. Senaratne DNS, Thakkar B, Smith BH, Hales TG, Marryat L, Colvin LA. The impact of adverse childhood experiences on multimorbidity: a systematic review and meta-analysis. BMC Med. 2024;22(1):315.

6. Finkelhor D, Ormrod RK, Turner HA. Poly-victimization: a neglected component in child victimization. Child Abuse Negl. 2007;31(1):7–26.

7. Lanza ST. Latent class analysis for developmental research. Child Dev Perspect. 2016;10(1):59–64.

8. Adams ZW, Moreland A, Cohen JR, Lee RC, Hanson RF, Danielson CK, et al. Polyvictimization: latent profiles and mental health outcomes in a clinical sample of adolescents. Psychol Violence. 2016;6(1):145–55.

9. Griffith EL, Ramarushton B, Davis KP, Contractor AA, Boals A. Relations between trauma-based subgroups and posttrauma health outcomes: a latent class analysis. Psychol Trauma. 2025;17(1):186–95.

10. Yapp E, Booth T, Davis K, Coleman J, Howard LM, Breen G, et al. Sex differences in experiences of multiple traumas and mental health problems in the UK Biobank cohort. Soc Psychiatry Psychiatr Epidemiol. 2023;58(12):1819–31.

11. Blosnich JR, Dichter ME, Cerulli C, Batten SV, Bossarte RM. Disparities in adverse childhood experiences among individuals with a history of military service. JAMA Psychiatry. 2014;71(9):1041–8.

12. MacManus D, Short R, Lane R, Jones M, Hull L, Howard LM, et al. Intimate partner violence and abuse experience and perpetration in UK military personnel compared to a general population cohort: a cross-sectional study. Lancet Reg Health Eur. 2022;20:100448.

13. Wilson LC. The prevalence of military sexual trauma: a meta-analysis. Trauma Violence Abuse. 2018;19(5):584–97.

14. Gibson CJ, Gray KE, Katon JG, Simpson TL, Lehavot K. Sexual assault, sexual harassment, and physical victimization during military service across age cohorts of women veterans. Womens Health Issues. 2016;26(2):225–31.

15. Campbell GM, Williamson V, Murphy D. “A Hidden Community”: The Experiences of Help-Seeking and Receiving Mental Health Treatment in U.K. Women Veterans. A Qualitative Study. Armed Forces & Society. 2023;51(1):22–45.

16. Jaycox LH, Morral AR, Street A, Schell TL, Kilpatrick D, Cottrell L. Gender differences in health among U.S. service members: unwanted gender-based experiences as an explanatory factor. RAND Health Q. 2023;10(2):8.

17. Montgomery AE, Cutuli JJ, Evans-Chase M, Treglia D, Culhane DP. Relationship among adverse childhood experiences, history of active military service, and adult outcomes: homelessness, mental health, and physical health. Am J Public Health. 2013;103 Suppl 2:S262–8.

18. Patten E, Parker K. Women in the U.S. military: growing share, distinctive profile. Washington, DC: Pew Research Center; 2011.

19. UK Biobank. UK Biobank: protocol for a large-scale prospective epidemiological resource. Stockport: UK Biobank; 2007.

20. Office for National Statistics. Standard Occupational Classification (SOC) 2010 [Internet]. Newport: ONS; 2016 [cited 2024 Mar]. Available from: https://www.ons.gov.uk/methodology/classificationsandstandards/standardoccupationalclassificationsoc/soc2010

21. Office for National Statistics. Standard Occupational Classification 2010: Volume 1, structure and descriptions of unit groups. Newport: ONS; 2010.

22. Grabe HJ, Schulz A, Schmidt CO, Appel K, Driessen M, Wingenfeld K, et al. [A brief instrument for the assessment of childhood abuse and neglect: the childhood trauma screener (CTS)]. Psychiatr Prax. 2012;39(3):109–15.

23. Kessler RC, Andrews G, Mroczek D, Ustun B, Wittchen H-U. The World Health Organization Composite International Diagnostic Interview short-form (CIDI-SF). Int J Methods Psychiatr Res. 1998;7(4):171–85.

24. World Health Organization. *International statistical classification of diseases and related health problems. 10th revision*. Geneva: World Health Organization; 2016.

25. Bush K, Kivlahan DR, McDonell MB, Fihn SD, Bradley KA. The AUDIT alcohol consumption questions (AUDIT-C): an effective brief screening test for problem drinking. Arch Intern Med. 1998;158(16):1789–95.

26. UK Ministry of Housing, Communities and Local Government. The English indices of deprivation 2019. London: Ministry of Housing, Communities and Local Government; 2019.

27. Katon JG, Lehavot K, Simpson TL, Williams EC, Barnett SB, Grossbard JR, et al. Adverse childhood experiences, military service, and adult health. Am J Prev Med. 2015;49(4):573–82.

28. Khalifeh H, Hargreaves J, Howard LM, Birdthistle I. Intimate partner violence and socioeconomic deprivation in England: findings from a national cross-sectional survey. Am J Public Health. 2013;103(3):462–72.

29. Juškaitė S, Stone J, Greenberg N, Dyball D, Fear NT. Current perspectives on the mental health of UK military personnel and veterans. Br Med Bull. 2025;154(1):ldaf003.

30. Larrazabal MA, Naragon-Gainey K, Conway CC. Distress tolerance and stress-induced emotion regulation behavior. J Res Pers. 2022;99:104243.

31. Kanesarajah J, Waller M, Zheng WY, Dobson AJ. Unit cohesion, traumatic exposure and mental health of military personnel. Occup Med (Lond). 2016;66(4):308–15.

